# Clinical, laboratory, and imaging findings of stage 3-5 chronic kidney disease patients suffering from COVID-19 in Bangladesh: a prospective cross-sectional study

**DOI:** 10.1101/2023.12.18.23300150

**Authors:** Ahsan Ullah, Asia Khanam, Mina Mondal, Md Rezwanul Haque, AHM Sanjedul Haque Sumon, Shakila Khan, Mohimanul Hoque, Motiur Rahman Sumon, Mohammad Meshbahur Rahman

## Abstract

**Background:** Chronic kidney disease (CKD) patients were susceptible to morbidity and mortality once they affected by COVID-19. These patients were more likely to develop severe disease, requiring dialysis, admission to intensive care unit. The aim of this study was to evaluate the presentations and outcomes of COVID-19 in stage 3-5 CKD patients not on dialysis.

**Methods:** This prospective observational study was conducted in the COVID-19 unit, at Bangabandhu Sheikh Mujib Medical University (BSMMU), Dhaka from September 2020 to August 2021. Hospitalized RT-PCR positive COVID-19 patients with pre-existing CKD having eGFR <60 ml/min/1.73 m^2^ but not yet on dialysis were enrolled. Clinical and laboratory parameters were recorded. Outcomes were observed till discharge from the hospital and followed up after 3 months of survived patients.

**Results:** Out of 109 patients, the mean age was 58.1(SD: ±15.4) years where 61.5% were male. Common co-morbid conditions were hypertension (89.0%), diabetes mellitus (58.7%) and ischemic heart disease (24.8%). Fever, cough, shortness of breath and fatigue were common presenting features. Most of the patients had moderate (41.3%) and severe (41.3%) COVID-19. Sixty-six patients (60.6%) developed AKI on CKD. Twenty patients (30.3%) required dialysis. Death occurred in 16 patients (14.7%) and 12 patient’s (11%) required ICU admission and 6 patients (9.1%) achieved baseline renal function at discharge. We identified risk factors like low haemoglobin, lymphopenia, high CRP, high procalcitonin, high LDH and low SpO_2_ in patients who did not survive. Seventy-six patients were followed up at 3rd month where 17 patients were lost. Ten patients (27.0%) achieved baseline renal function who had persistent AKI at discharge and 34 patients (87.1%) remained stable who had stable renal function at discharge.

**Conclusion:** The stage 3-5 chronic kidney patients with COVID-19 are vulnerable to severe to critical morbidity and mortality with higher incidence of AKI which demands a special attention to this group of patients.

## Introduction

The COVID-19 pandemic caused by the SARS-CoV-2 virus has emerged as a global public health emergency, with millions of reported cases and fatalities worldwide [1–7]. Chronic kidney disease (CKD) is a common comorbidity among COVID-19 patients, which is associated with increased morbidity and mortality [8–10]. Although most patients with COVID-19 develop mild symptoms, those with comorbidities such as chronic kidney disease (CKD) are at increased risk of severe disease and death [11]. Patients with CKD are not only at a higher risk of infection, but also prone to severe complications once infected because of their immunocompromised status [12]. Several studies have demonstrated that patients with CKD have a higher risk of mortality, require dialysis, admission to intensive care units (ICUs), mechanical ventilation, and are more likely to die than those without CKD [9, 11, 13–18]. Stage 3-5 CKD patients are particularly vulnerable due to their impaired immune system and pre-existing comorbidities [19, 20].

Globally, several studies have reported that CKD is associated with a higher risk of severe COVID-19 and mortality [12–14, 21–27]. According to the World Health Organization (WHO), as of 19 April 2023, over 763.74 million confirmed cases and 6.91 million deaths have been reported worldwide due to COVID-19 [28]. Among COVID-19 patients, the prevalence of CKD varies between 4.4% to 33%, depending on the population studied and the definition of CKD used [9, 14, 15]. In Bangladesh, there has been a significant increase in the number of COVID-19 cases since the first case was reported in March 2020 [1, 3]. The prevalence of CKD among COVID-19 patients in Bangladesh is not well-documented, but it is estimated that a significant proportion of COVID-19 patients have underlying CKD.

CKD patients with COVID-19 may present with atypical symptoms or have a more severe disease course compared to non-CKD patients [29, 30]. Common symptoms reported in CKD patients with COVID-19 include fever, cough, dyspnea, fatigue, and gastrointestinal symptoms [25, 26]. Cardiovascular complications are also more common in CKD patients with COVID-19, such as myocardial injury, arrhythmias, and heart failure [23, 31]. Laboratory findings in CKD patients with COVID-19 may include elevated inflammatory markers such as C-reactive protein (CRP), ferritin, and interleukin-6 (IL-6), as well as lymphopenia, thrombocytopenia, and elevated creatinine [9]. Imaging studies such as chest X-ray and computed tomography (CT) scans may show characteristic features of COVID-19 pneumonia, such as ground-glass opacities, consolidation, and interstitial infiltrates [26, 32].

CKD patients with COVID-19 have been reported to have a higher risk of severe disease, need for mechanical ventilation, and mortality compared to non-CKD patients. In a study, CKD was associated with a 3-fold higher risk of mortality among COVID-19 patients [12]. Moreover, the severity of CKD is also associated with worse outcomes, with stage 5 CKD patients having the highest mortality risk [19, 20]. An analysis of 7162 laboratory-confirmed COVID-19 cases in the USA demonstrated that CKD was 12-fold more frequent in those with ICU admission and 9-fold more frequent in hospitalized, non-ICU COVID-19 patients than in those not hospitalized [12].

Patients with underlying conditions such as diabetes, cardiovascular disease, liver cirrhosis, and CKD have a higher risk of infection. Patients with pre-existing CKD have elevated levels of C-reactive protein, ferritin, and lactate dehydrogenase compared to patients without CKD [9]. They are more likely to progress to severe disease, requiring admission to the ICU and mechanical ventilation leading to death. Although the incidence of ICU admission ranges from 11.4-23.5%, (95%CI: 0.114-0.235), the risk of mortality can vary between 20.5-26.1% [18].

Bangladesh is one of the countries affected by the COVID-19 pandemic, with a significant burden of CKD patients[33, 34]. The outcome of CKD patients with COVID-19 is not well-documented here. However, the country has a high burden of CKD patients, and COVID-19 may exacerbate the disease, leading to a higher risk of morbidity and mortality [34]. Given the increased risk of severe disease in CKD patients with COVID-19, there is a need to evaluate their clinical, laboratory, and imaging findings, as well as disease outcome. Considering this, this study aims to provide a comprehensive analysis of the clinical, laboratory, and imaging findings, as well as disease outcome of stage 3-5 CKD patients with COVID-19. The findings of this study will provide valuable insights into the management of this vulnerable patient population during the current pandemic and inform future research directions.

## Methods

### Ethical permission and consent for data collection

Prior to commencement of this study, the research protocol was approved by the Institutional Review Board (IRB) of Bangabandhu Sheikh Mujib Medical University (BSMMU), Dhaka, Bangladesh with the IRB number BSMMU/2020/9625. The aims and objectives of the study along with its procedure, risk and benefits of this study was explained to the patients/guardian in easily understandable local language and then informed consent had taken from each patient or guardian (if patient is incapacitated/incompetent). It was assured that all information and records will be kept confidential. Participants reserved the rights of withdrawal during any time of thesis work.

### Study setting

This prospective cross-sectional study was conducted at the COVID-19 dedicated unit of Bangabandhu Sheikh Mujib Medical University (BSMMU), Dhaka, Bangladesh from September 2020 to August 2021.

### Study population

Patients who were admitted in COVID unit of Bangabandhu Sheikh Mujib Medical University, Dhaka, Bangladesh, were laboratory-confirmed SARS-CoV-2 infection by real-time reverse transcription-polymerase chain reaction (RT-PCR) and who had pre-existing chronic kidney disease fulfilling inclusion and exclusion criteria were taken as the study population. Follow-up was conducted after three months after at the Department of Nephrology, BSMMU, Dhaka.

### Sampling and sample size

Purposive sampling technique was applied where the sample size was determined by the formula 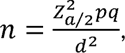 Where, *Z*_*a*/2_ = 1.96 for 5% level of significance, p= prevalence of CKD occurrence with COVID-19= 0.116, q=1-p=0.884, d= level of absolute precision=0.06 [29]. Applying the formula, the final sample size became 109.

### Selection criterion

The study had some selection criterion. The inclusion criteria were i) confirmed diagnosis of COVID-19 patients through real-time reverse transcriptase polymerase chain reaction (RT-PCR), ii) confirmed diagnosis of CKD patients with eGFR <60 ml/min/1.73 m^2^ based on kidney disease: Improving Global Outcomes (KDIGO) criteria; and iii) aged older than 18 years. The exclusion criterion was i) CKD patients on maintenance haemodialysis and ii) renal allograft recipient patients.

### Study variables

The variables included in the study were the patients’ demographic variables (e.g., age, sex, BMI, comorbidities, Duration of CKD, Duration from onset of illness to admission, Clinical presentation of COVID-19, Oxygen saturation (SpO_2_) (%) at admission), Laboratory and imaging variables (e.g., Complete blood count, Serum creatinine, Serum electrolytes, Urine R/M/E, CRP, Procalcitonin, Ferritin, D-dimer, LDH, HRCT scan of chest), and the outcome variables (e.g., length of hospital stay, outcome at discharge, AKI on CKD, death, underwent dialysis and recovered to baseline renal function).

### Data collection procedure

After getting IRB approval, patients were recruited from the dedicated COVID-19 unit of Bangabandhu Sheikh Mujib Medical University (BSMMU), Dhaka. Subjects were selected on the basis of enrolment criteria. Patients of CKD was defined as the presence of decreased kidney function (eGFR <60 ml/min/1.73 m^2^) from their previous medical record for more than three months. COVID-19 positive patients were confirmed by RT-PCR test from nasopharyngeal or oropharyngeal swab. According to the institutional protocol of BSMMU, the disease severity was classified into-mild, moderate, severe and critical. The data were collected through a pre-designed questionnaire after taking consent. Initial information of the patients was collected form hospital record or treatment files. Then the history, clinical examination and laboratory investigation reports were obtained from each patient. All laboratory investigation were done in the emergency COVID-19 laboratory and laboratory service centre of BSMMU. The serum creatinine was recorded at 0, 48^th^ hour and 7^th^ day and at discharge. Subjects were followed-up until discharged from hospital or death. Outcome variables were measured as death during hospital stay, length of hospital stays, need for dialysis, recovery of renal function during hospital stay and persistent AKI during discharge from hospital. At the 3^rd^ month survived patients were followed up for persistence of symptom and outcome measured as death, need for dialysis and recovery of renal function.

### Statistical analysis

All data were recorded systematically in preformed data collection form. The descriptive analysis was performed first to present the demographic, clinical and laboratory findings. The difference in means or percentages of different variables were performed using either the chi-square test for nominal variables or the student t-test for numerical variables. Statistical analyses were performed by using windows-based computer software with Statistical Packages for Social Sciences (SPSS-26) (SPSS Inc, Chicago, IL, USA). For the analysis of outcomes, p-values less than 0.05 was considered as significant.

## Results

### Demographic characteristics of patients

Among 109 stage 3-5 CKD patients with COVID-19, the mean age of the respondents was 58.1 (SD: ±15.4) years with age range 25-85 years. About 23.9% of patients were in the age group 51-60 years following 19.3% and 17.4% were in the age group 61-70 years and 71-80 years respectively. Sixty-seven patients (61.5%) were male and the rest were female. The average BMI was 24.54 (18.50-30.50) kg/m^2^ which indicates overweight and obesity status of the respondents (Table 1).

**Table 1.**
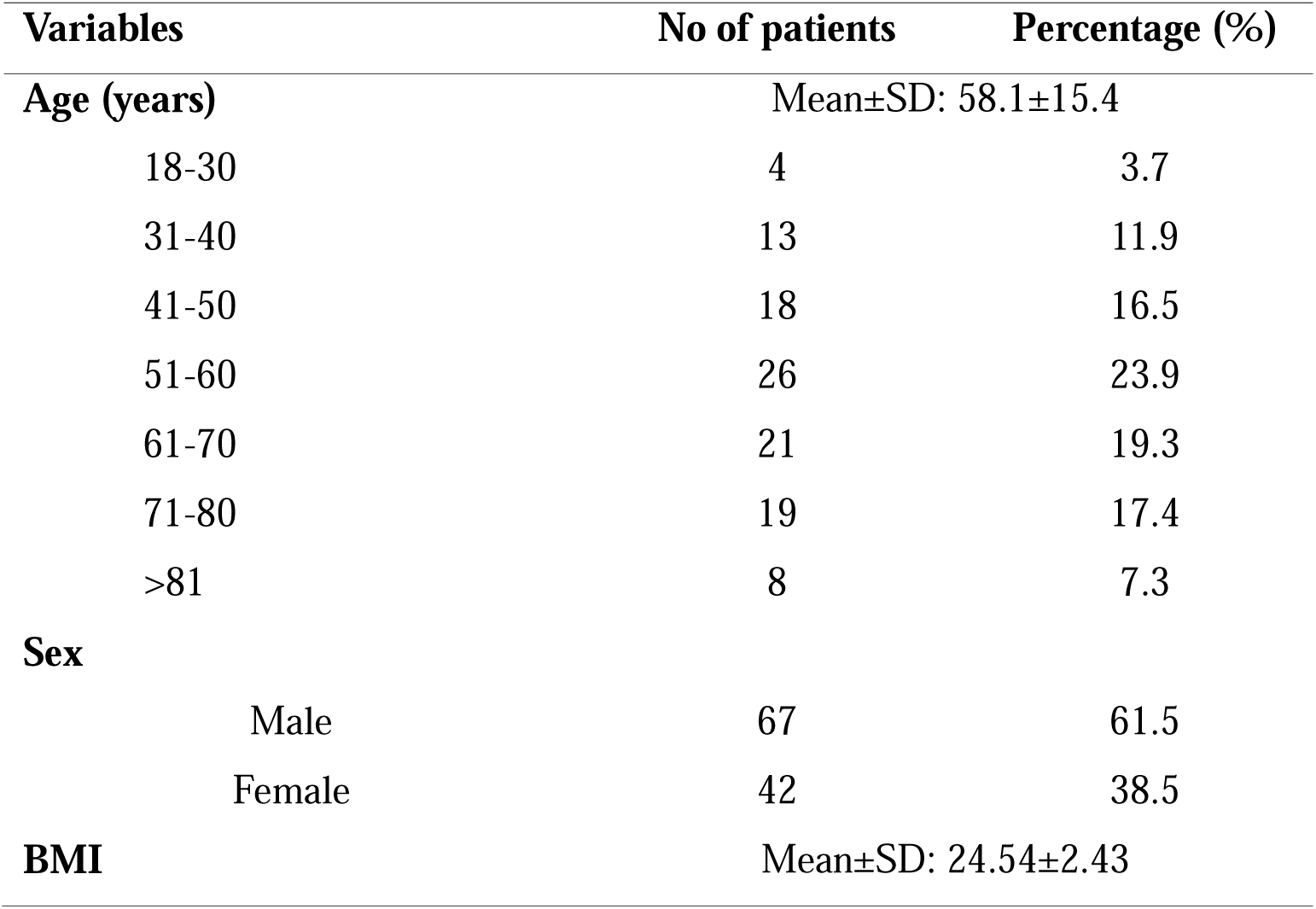
Demographic characteristics of the study subjects (n=109).

### Clinical presentation of stage 3-5 CKD patients with COVID-19

Clinical characteristics including comorbidities of 109 stage 3-5 CKD patients with COVID-19 were investigated (Table 2). Among the patients, majority of them were suffering from Fever (74.3%), following 70.6% had cough, 67% had shortness of breath, 51.4% had fatigue, and 16.5% had Diarrhoea. The mean SpO_2_ was 90.1±7.22 and the average respiratory rate was high (26.16±4.26).

**Table 2.** Clinical presentation of the patients (n=109).

| Variables | No of patients | Percentage | Mean±SD |
| --- | --- | --- | --- |
| **Symptoms at diagnosis** |  |  |  |
| Fever | 81 | 74.3 |  |
| Cough | 77 | 70.6 |  |
| Shortness of breath | 73 | 67.0 |  |
| Diarrhoea | 18 | 16.5 |  |
| Fatigue | 56 | 51.4 |  |
| SpO<sub>2</sub> | 109 | 100.0 | 90.1±7.22 |
| Respiratory rate | 109 | 100.0 | 26.16±4.26 |
| **Comorbidities** |  |  |  |
| Diabetes mellitus | 64 | 58.7 |  |
| Hypertension | 97 | 89.0 |  |
| Ischemic heart disease | 27 | 24.8 |  |
| Cerebrovascular disease | 9 | 8.3 |  |
| Obstructive airway disease | 24 | 22.0 |  |
| **HRCT of Chest for Lungs involvement** |  |  |  |
| <25% | 16 | 14.68 |  |
| 26-50% | 73 | 66.97 |  |
| >50% | 8 | 7.34 |  |
| >75 | 12 | 11.01 |  |

Hypertension (89%) was the most common comorbidity among the stage 3-5 CKD patients who were suffering from COVID-19. Others comorbidities were diabetes mellitus (58.7%), ischemic heart disease (24.8%), obstructive airway disease (22.0%) and cerebrovascular disease (8.3%). About 67% patients had 26-50% Lung involvement found in HRCT scan of the chest (Table 2).

### Patients’ severity in COVID-19

Most of the stage 3-5 CKD patients were suffering from moderate (41.28%) and severe (41.28%) COVID-19. Although, females were found more (14.30% versus 9.0%) critical condition, the severity percentage was higher (46.3% versus 33.30%) among male than female (Figure 1).

**Figure 1.**
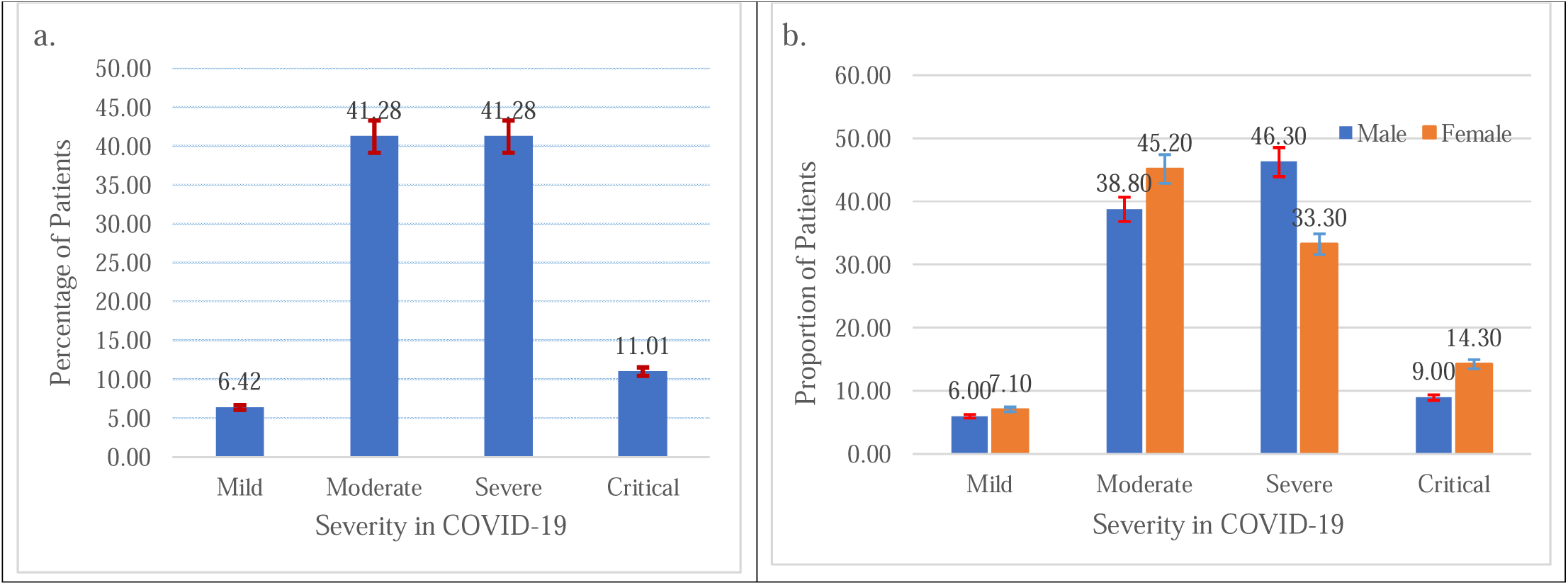
Severity of stage 3-5 CKD patients who were suffering from COVID-19-a. overall severity; b. severity by Sex of the patients.

### Laboratory findings of stage 3-5 CKD patients suffering from COVID-19

Patients’ laboratory findings were presented as mean, standard deviation and range. The mean haemoglobin was 9.85±1.55 gm/dl, lymphocyte was 17.03±5.18 %. The mean baseline serum creatinine was 3.99±1.70 mg/dl, Sodium level was 131.61±7.50 mmol/l, Ferritin was 754.45±414.88 ng/ml, D-dimer 1.05±1.00 µg/ml, CRP 83.23±42.21 mg/l, Procalcitonin 1.52±1.41 ng/ml and LDH was 454.14±214.06 u/l (Table 3).

**Table 3.** Laboratory findings of the study subjects (n=109)

| Variables | Mean ±SD (Ranges) |
| --- | --- |
| Hb (gm/dl) | $9.85 \pm 1.55$ (7.0-13.60) |
| Lymphocyte (%) | $17.03 \pm 5.18$ (8.0-29.0) |
| Serum creatinine (mg/dl) | $3.99 \pm 1.70$ (1.30-7.30) |
| Sodium (mmol/l) | $131.61 \pm 7.50$ (112.0-155.0) |
| Ferritin (ng/ml) | 754.45±314.88 (101.0-3047.0) |
| D dimer (mg/l) | 1.05±1.00 (0.05-4.50) |
| CRP (mg/l) | 83.23±42.21 (10.0-210.0) |
| Procalcitonin (ng/ml) | 1.52±1.41 (0.05-8.83) |
| LDH (u/l) | 454.14±214.06 (216.0-960.0) |

### Distribution of stage 3-5 CKD patients by in-hospital outcomes

Sixty-six (60.6%) patients developed acute kidney injury of which 20 (30.3%) patient required dialysis, 12 (11.0%) patient needed ICU admission. Six (9.1%) patients recovered to baseline renal function. We observed 16 (14.7%) deaths out of 109 patients. Average length of hospital stay was 13.71±5.66 days (Table 4).

**Table 4.** Distribution of the stage 3-5 CKD patients by in-hospital outcomes (n=109)

| Variables |  | No of patients | Percentage (%) |
| --- | --- | --- | --- |
| AKI on CKD |  | 66 | 60.6 |
| Requiring dialysis |  | 20 | 30.3 |
| Recovery of renal function to baseline |  | 6 | 9.1 |
| Discharged with persistent AKI |  | 44 | 66.6 |
| ICU admission |  | 12 | 11.0 |
| Death |  | 16 | 14.7 |
| Length of hospital stay (days) | Mean±SD | 13.71±5.66 |  |
|  | Range | (2-25) days |  |

### Comparison of baseline characteristics between AKI on CKD and Stable CKD patients

Comparison of baseline characteristics between AKI on CKD and Stable CKD revealed that patients who developed AKI had more delay in admission from symptom onset (8.93±2.05 vs 6.83±1.66, p=<0.001). Patients were aged in AKI group and had higher incidence of co-morbidities than stable CKD patients but not statistically significant (Table 5).

**Table 5.**
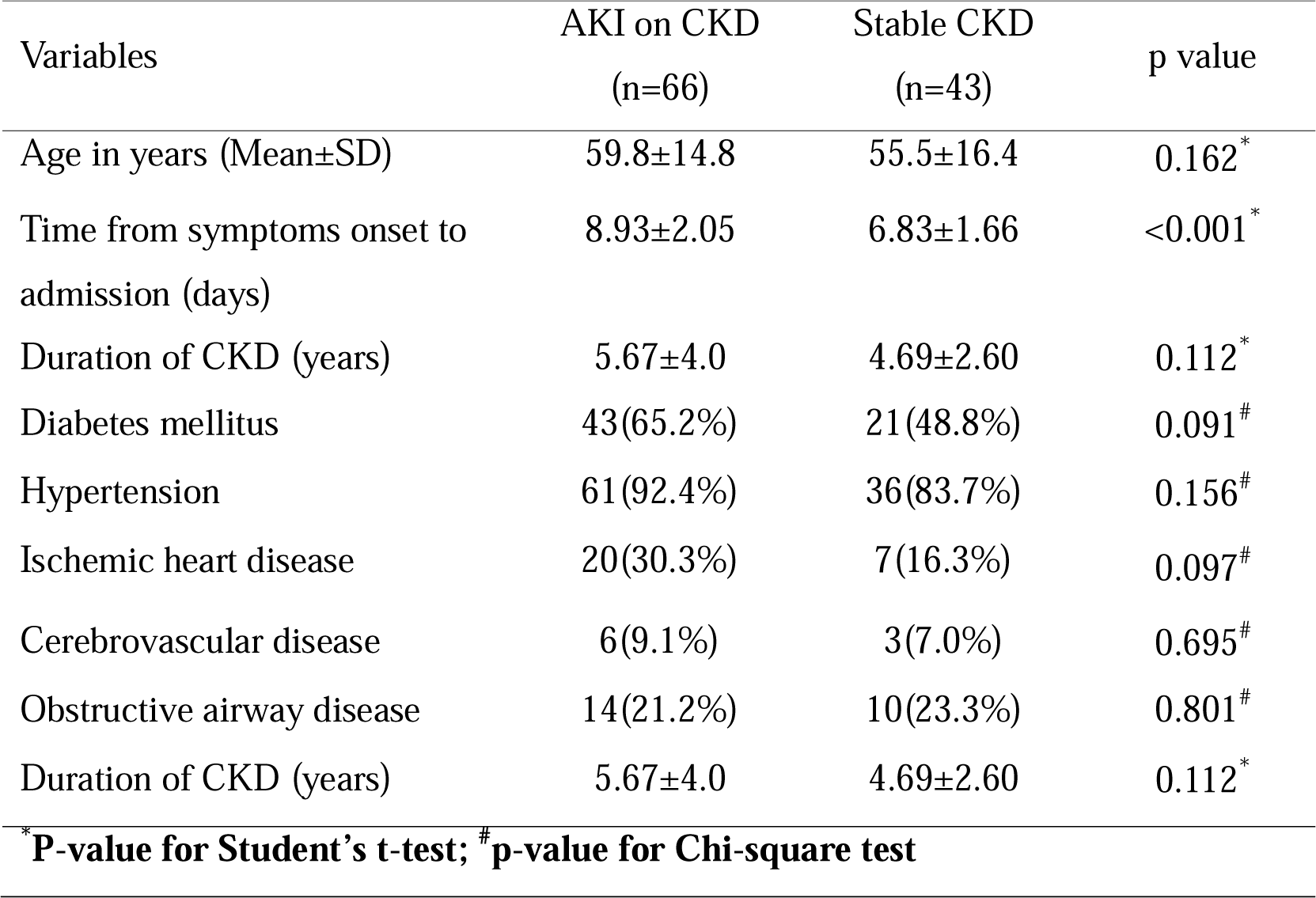
Comparison of baseline characteristics between AKI on CKD and Stable CKD (N=109)

### Comparison of laboratory findings between AKI on CKD and stable CKD patients

Comparison of laboratory parameters between AKI on CKD and stable CKD showed hyponatremia (129.71±8.33 vs 134.53±4.76, p=0.001) significantly higher in stable CKD patients. The rest laboratory parameters were found significantly higher in patients with acute kidney infection (Table 6).

**Table 6.**
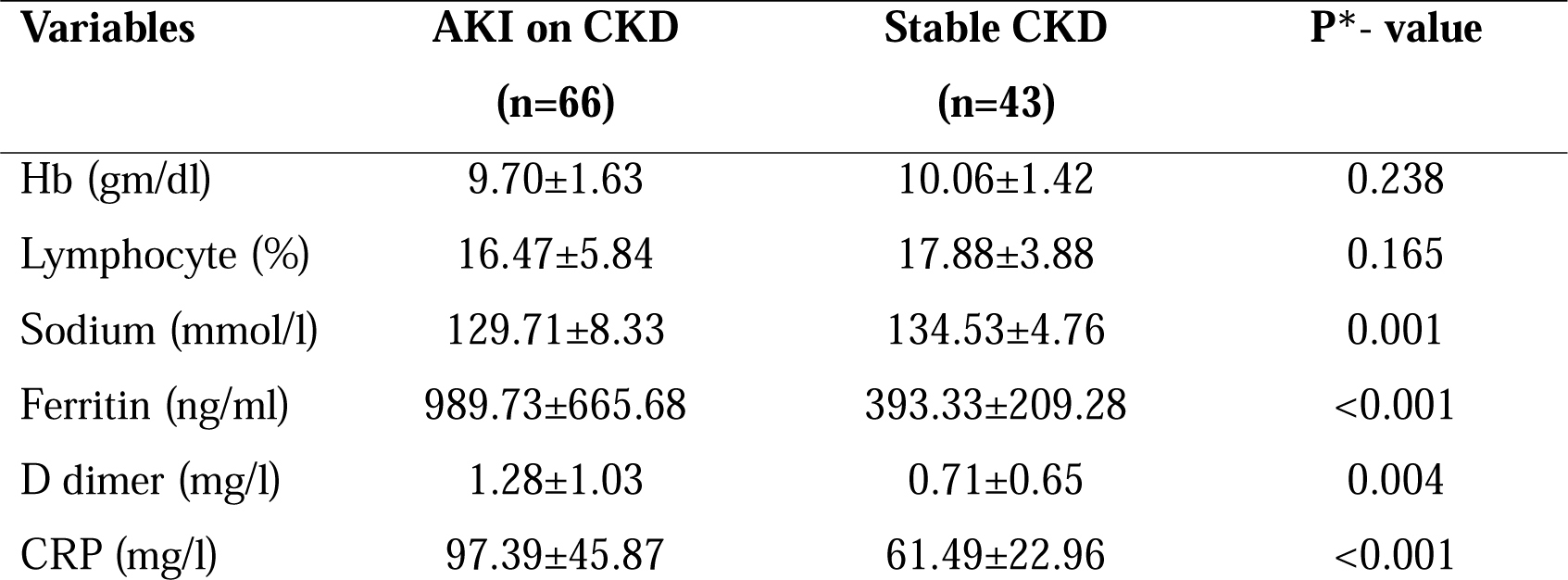
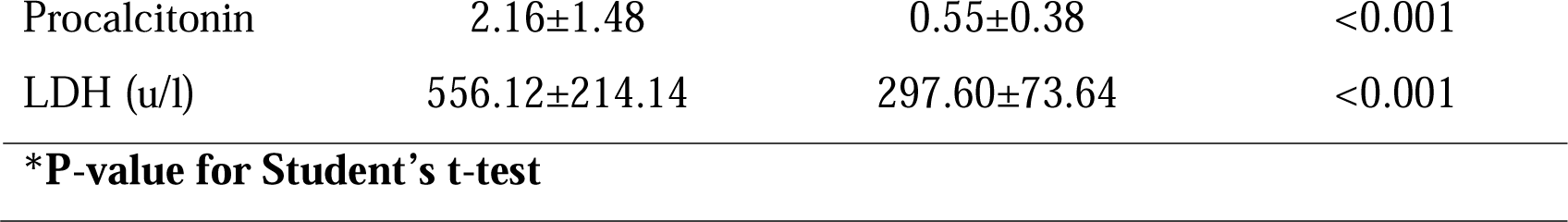
Comparison of laboratory findings between AKI on CKD and stable CKD (n=109)

### Comparison of laboratory findings between survived and non- survived patients

Comparison of laboratory parameters showed hemoglobin (8.90±1.17 vs 10.01±1.56, p=0.008), lymphopenia (13.81±4.23 vs 17.58±5.15, p=0.007), CRP (103.88±33.89 vs 79.67±42.63, p=0.033), LDH (594.50±177.26 vs 429.99±211.30, P=0.004) were significantly higher among non-survivor patients (Table 7). On the other hand, AKI on CKD (100.0% vs 53.8%, p=<0.001), requiring dialysis (50.0% vs 12.9%, p<0.001), ICU admission (50.0% vs 4.3%, p<0.001) were higher in non-survivors than survived patient (Table 7).

**Table 7.** Comparison of laboratory findings and SpO_2_ between survived and non- survived subjects (n=109)

| Variables | Non-survivors<br>(n=16) | Survivors<br>(n=93) | p value |
| --- | --- | --- | --- |
| Hb(gm/dl) | 8.90±1.17 | 10.01±1.56 | 0.008<sup>*</sup> |
| Lymphocyte (%) | 13.81±4.23 | 17.58±5.15 | 0.007<sup>*</sup> |
| Serum creatinine at admission(mg/dl) | 4.28±1.91 | 3.44±1.63 | 0.067<sup>*</sup> |
| Ferritin(ng/ml) | 718.25±367.88 | 760.68±370.17 | 0.848<sup>*</sup> |
| D dimer(mg/l) | 1.31±1.1 | 1.01±0.73 | 0.279<sup>*</sup> |
| CRP (mg/l) | 103.88±33.89 | 79.67±42.63 | 0.033<sup>*</sup> |
| Procalcitonin (ng/ml) | 3.42±0.83 | 1.19±1.22 | 0.339<sup>*</sup> |
| LDH(u/l) | 594.50±177.26 | 429.99±211.30 | 0.004<sup>*</sup> |
| SpO<sub>2</sub> (%) | 84.0±4.7 | 91.1±7.1 | <0.001<sup>#</sup> |
| AKI on CKD | 16(100.0%) | 50(53.8%) | <0.001<sup>#</sup> |
| Requiring dialysis | 8(50.0%) | 12(12.9%) | <0.001<sup>#</sup> |
| ICU admission | 8(50.0%) | 4(4.3%) | <0.001<sup>#</sup> |
| **\*P-value for Student's t-test; <sup>#</sup>p-value for Chi-square test** |  |  |  |

### Comparison of COVID-19 severity and other outcomes among CKD stages

Comparison of patients’ severity among different CKD stages found that CKD stage 4 and 5 had significantly (p = 0.001) more severe to critical COVID-19 compared to stage 3 CKD patients. The frequency of AKI was significantly (p=0.005) higher along with the advancement of CKD stage. Stage 5 CKD patients needed significantly more dialysis (75.0%) and ICU admission (83.3%) compared to other stage. Death occurred significantly (p = 0.024) higher in stage 5 CKD (62.5%) compared to stage 3 and stage 4 (Table 8).

**Table 8.**
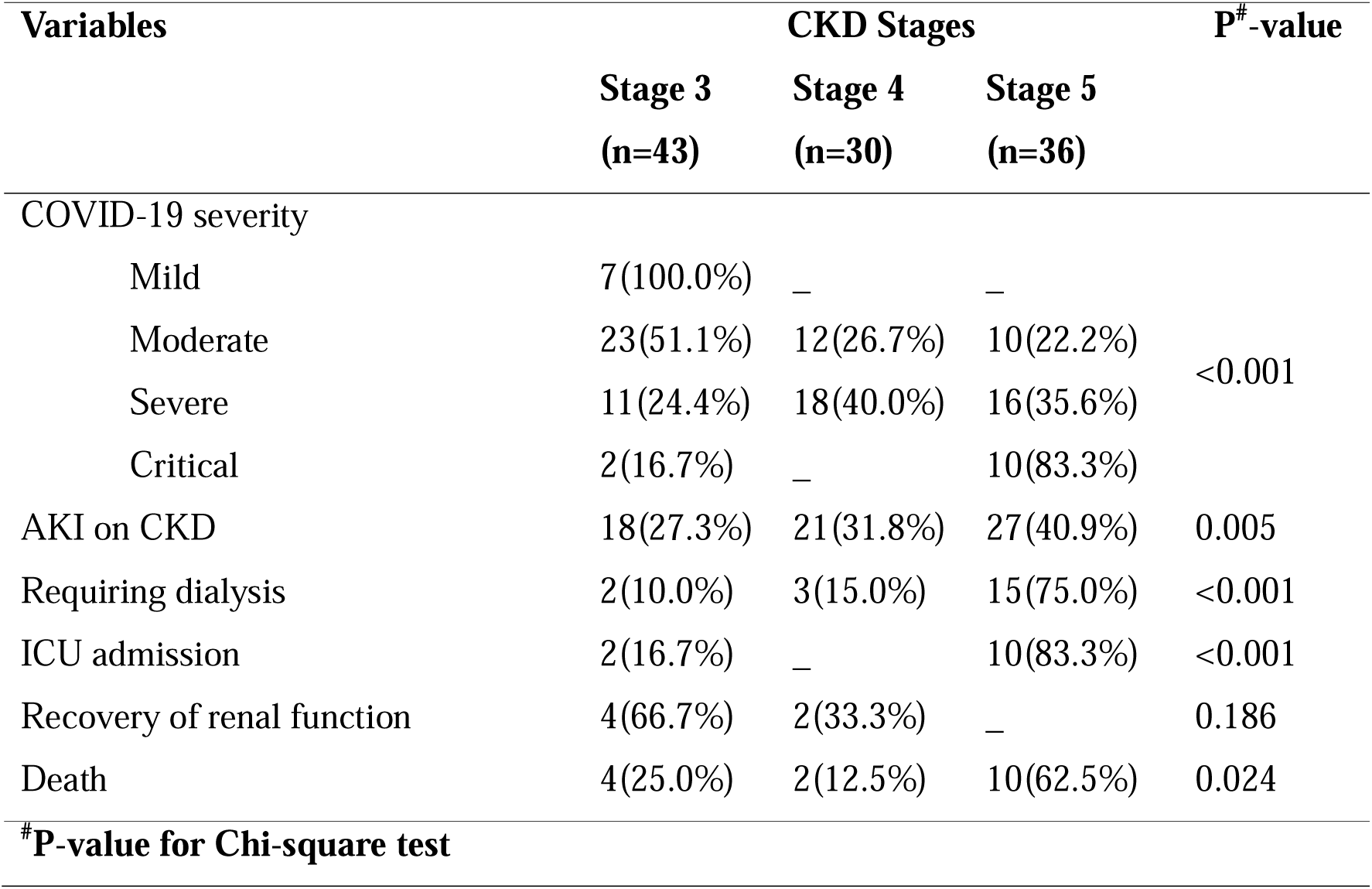
Comparison of COVID-19 severity and outcomes among CKD stages (n=109)

| Variables | CKD Stages |  |  | P<sup>#</sup>-value |
| --- | --- | --- | --- | --- |
|  | Stage 3 | Stage 4 | Stage 5 |  |
|  | (n=43) | (n=30) | (n=36) |  |
| COVID-19 severity |  |  |  |  |
| Mild | 7(100.0%) | — | — | <0.001 |
| Moderate | 23(51.1%) | 12(26.7%) | 10(22.2%) |  |
| Severe | 11(24.4%) | 18(40.0%) | 16(35.6%) |  |
| Critical | 2(16.7%) | — | 10(83.3%) |  |
| AKI on CKD | 18(27.3%) | 21(31.8%) | 27(40.9%) | 0.005 |
| Requiring dialysis | 2(10.0%) | 3(15.0%) | 15(75.0%) | <0.001 |
| ICU admission | 2(16.7%) | — | 10(83.3%) | <0.001 |
| Recovery of renal function | 4(66.7%) | 2(33.3%) | — | 0.186 |
| Death | 4(25.0%) | 2(12.5%) | 10(62.5%) | 0.024 |
| <sup>#</sup>P-value for Chi-square test |  |  |  |  |

### Findings after three months follow up

After three months, 17 (15.59%) patients had lost to follow up, and fatigue (65.8%) and cough (38.2%) had the most common symptoms among the rest patients. Six patients (7.9%) had shortness of breath and 4 patients (5.3%) had persistent anosmia (Supplementary Table S1). During discharge, 44 patients had persistent AKI. Out of them, 37 patients reported at follow up. At last month to follow up, 25 patients (67.5%) did not achieved recovery to baseline renal function, among them 11 patients (29.7%) required dialysis, 10 patients (27.0%) recovered to baseline renal function and death occurred in 2 patients (Supplementary Table S2). During discharge, 49 patients had persistent stable renal function. Follow up done on 39 patients where 10 patients again lost to follow up. At third month to follow up, five patients (12.8%) developed AKI, and among them 01 patients required dialysis. About 34 (87.1%) patients had stable renal function (Supplementary Table S3 and S4).

## Discussion

The findings of the study highlight the clinical and laboratory characteristics of stage 3 to 5 chronic kidney disease (CKD) patients with COVID-19. This prospective cross-sectional study included 109 patients with a mean age of 58.1 years, suggesting that elderly patients were more commonly affected. In terms of gender, about 61.5% of the patients were male which is consistent with previous research that has recommended that males may be at higher risk for severe COVID-19 outcomes, including hospitalization and mortality [4, 10, 35, 36]. Obesity has been identified as a risk factor for severe COVID-19 outcomes, including increased hospitalization rates and worse clinical outcomes [10, 35, 37]. In this study, the average body mass index (BMI) of the patients was 24.54 kg/m^2^, indicating the majority patients were overweight and obese on average. Therefore, the high prevalence of overweight and obesity among the CKD patients in this study may be an important consideration in managing their risk for COVID-19 complications.

The majority of these patients presented with fever (74.3%), followed by cough (70.6%), shortness of breath (67%), fatigue (51.4%), and diarrhea (16.5%). The mean oxygen saturation (SpO2) was 90.1±7.22, indicating a lower-than-normal level, and the average respiratory rate was high at 26.16±4.26 breaths per minute. Literature suggests that the observed high symptoms and clinical parameters among the patients are likely due to the combined effects of the viral infection and pre-existing kidney disease [10, 38]. Fever and cough are common in viral infections; shortness of breath and compromised oxygen saturation result from respiratory involvement, exacerbated by compromised kidney and lung function; and fatigue may be heightened due to the interaction of both conditions are reported in COVID-19 [39, 40].

Notably, about 67% of the patients had 26-50% lung involvement which highlight the significant burden of comorbidities in CKD patients with COVID-19. One or more comorbid conditions including hypertension and diabetes mellitus are the prevalent in stage 3 to 5 COVID-19 patients who are suffering from COVID-19, and the findings align with the current study [41–43]. The high percentage of lung involvement in HRCT scans suggests that CKD patients may be at increased risk of respiratory complications when infected with COVID-19 [38]. The findings of this study emphasize the importance of closely monitoring CKD patients with COVID-19, particularly those with comorbidities, for early detection and management of respiratory complications [44]. It is crucial to implement appropriate preventive measures, such as regular monitoring of oxygen saturation levels and respiratory rates, as well as optimal management of comorbidities, to minimize the morbidity and mortality associated with COVID-19 in CKD patients [45].

After the three months follow-up, the study findings reveal concerning outcomes with 5.4% succumbing to mortality. For those with stable renal function during discharge, 12.8% developed AKI after three months, with 87.1% maintaining stability. The findings underscore the importance of intensive care, targeted interventions, and improved follow-up strategies for CKD patients recovering from COVID-19, emphasizing the severity and long-term consequences of the virus in this vulnerable population [45–47]. The study also suggests that CKD patients, particularly those in advanced stages, are susceptible to moderate to severe COVID-19 illness that are comparable to existing literature [48–52]. It is important to note that CKD patients, regardless of gender, should be considered a high-risk group for COVID-19 due to their compromised immune function and underlying health conditions. Close monitoring of CKD patients, timely interventions, and appropriate management strategies should be implemented to minimize the severity of COVID-19 in this vulnerable population.

## Conclusion

The findings of the study emphasize the increased vulnerability of stage 3-5 CKD patients to moderate to severe COVID-19 illness, particularly in those with comorbidities such as hypertension and diabetes. The observed gender differences in the severity of COVID-19 among CKD patients, with a higher proportion of females in critical condition but higher severity percentage among males, warrant further investigation to better understand the underlying reasons and inform tailored interventions. The findings also highlight the need for implementing appropriate preventive measures, optimizing management of comorbidities, and close monitoring of respiratory parameters in CKD patients with COVID-19 to minimize the morbidity and mortality associated with the disease.

## Acknowledgements

The National Institute of Preventive and Social Medicine (NIPSOM) is an apex public health education and research institution in Bangladesh. We thank the Department of Biostatistics, NIPSOM for technically supporting the study for in analysis. We are also thankful to the participants who provided time and shared their health experiences.

## Data availability statement

The datasets generated and/or analyzed during the current study are available from the corresponding author on reasonable request to.

