## Supplemental Table 1-4 for "Clinical, laboratory, and imaging findings of stage 3-5 chronic kidney disease patients suffering from COVID-19 in Bangladesh: a prospective cross-sectional study"

**Supplementary files**

Table S1. Symptoms persist after 3 months (n=76)

| **Symptoms persist at 3^rd^ months** | **No of patients** | **Percentage** |
| --- | --- | --- |
| Fatigue | 50 | 65.8 |
| Cough | 29 | 38.2 |
| Shortness of breath | 6 | 7.9 |
| Anosmia | 4 | 5.3 |

Table S2. Outcome after 3 months who had persistent deterioration of renal function at discharge (n=37)

| **Variables** | **No of patients** | **Percentage** |
| --- | --- | --- |
| Not recovered to baseline renal function | 25 | 67.5% |
| Recovered to baseline renal function | 10 | 27.0% |
| Requiring dialysis | 11 | 29.7% |
| Death | 02 | 5.4% |

Table S3. Comparison of baseline parameters in relation to renal function recovery after 3 months

| **Variables** | **Not recovered**  **(n=25)** | **Recovered**  **(n=10)** | **p value** |
| --- | --- | --- | --- |
| Hb (gm/dl) | 9.53±1.35 | 9.89±1.40 | 0.488 |
| Lymphocyte (%) | 15.44±5.35 | 19.20±5.18 | 0.067 |
| Ferritin (ng/ml) | 1206.04±1058.15 | 760.00±355.40 | 0.205 |
| D dimer (mg/l) | 1.20±0.92 | 1.51±1.21 | 0.421 |
| S. creatinine (mg/dl) | 4.25±1.63 | 3.10±1.20 | 0.051 |
| CRP (mg/l) | 120.72±44.68 | 80.48±34.79 | 0.016 |
| Procalcitonin | 2.22±1.67 | 0.84±0.64 | 0.016 |
| LDH (u/l) | 649.20±223.48 | 433.70±155.58 | 0.009 |
| Duration of CKD (years) | 5.48±3.06 | 4.82±3.27 | 0.575 |
| COVID-19 severity |  |  |  |
| Moderate | 6(24.0%) | 6(60.0%) |  |
| Severe | 15(60.0%) | 4(40.0%) | 0.090 |
| Critical | 4(16.0%) | - |  |

Table S4. Outcome at 3^rd^ month who had of stable CKD at discharge (n=39)

| **Variables** | **No of patients** | **Percentage** |
| --- | --- | --- |
| Stable renal function | 34 | 87.1% |
| AKI on CKD | 05 | 12.8% |
| Requiring dialysis | 01 | 2.5% |
